# Association of Brain Structural Measurements and Polygenic Risk Scores with Obsessive-Compulsive Symptoms in Adolescents Diagnosed with Obsessive-Compulsive Disorder, Attention-Deficit/Hyperactivity Disorder, Anxiety, Depression, Autism and Tic Disorders

**DOI:** 10.64898/2025.12.17.25342484

**Authors:** Lilit Antonyan, S-M Shaheen, Christie L Burton, Gregory Baldwin, Rebecca Neill, Phillip Easter, Frank MacMaster, Gregory L. Hanna, David Rosenberg, Paul D. Arnold

**Affiliations:** The Mathison Centre for Mental Health Research & Education, Hotchkiss Brain Institute, University of Calgary, Calgary AB, Canada; Hospital for Sick Children, Toronto, ON, Canada; Department of Psychiatry & Behavioral Neurosciences, Wayne State University, Detroit MI, USA; IWK Health, Dalhousie University, Halifax NS, Canada; Department of Psychiatry, University of Michigan, Ann Arbor MI, USA

## Abstract

Obsessive-compulsive symptoms, characterized by intrusive thoughts and repetitive behaviors, are prevalent among youth. These symptoms are known to be moderately heritable and linked to structural brain changes involved in their pathophysiology. This study investigates the connections between structural brain alterations (cortical thickness, surface area and subcortical volume), genetic variation, and childhood obsessive-compulsive symptom scores within 143 samples of healthy control participants and cases diagnosed with obsessive-compulsive disorder, attention-deficit/hyperactivity disorder, anxiety disorder, autism spectrum disorders and/or tic disorders.

We hypothesize that the effect of genetic variants on standardized scores of obsessive-compulsive symptoms is mediated by imaging endophenotypes. To do so we test for associations between polygenic risk scores and structural imaging phenotypes within cortico-striato-thalamo-cortical circuitry and perform mendelian randomization analyses to identify potential causal pathways linking polygenic risk scores of structural brain alterations and obsessive-compulsive symptoms assessed with the Obsessive-Compulsive Subscale of the Child Behavior Checklist.

We observed that changes in cortical thickness of rostral middle frontal cortex and surface area of orbitofrontal cortex, along with other four regions have a significant genetic contribution in obsessive-compulsive symptom severity in adolescent samples. Additionally, surface area of inferior parietal lobule may act as a causal mediator between high-risk variants and obsessive-compulsive symptoms. The mentioned three regions are part of cortico-striato-thalamo-cortical circuitry that have various regulatory effects on obsessive-compulsive symptoms.

If these findings replicated in larger samples, they could offer valuable insights into the neurobiology of obsessive-compulsive traits and related structural alterations in specific brain regions.

## Introduction

Obsessive-compulsive (OC) symptoms (OCS) are core features of obsessive-compulsive disorder (OCD) and frequently co-occur with other psychiatric disorders in youth, including tic disorders (TD), anxiety disorders (ANX), autism spectrum disorders (ASD) and others. Indeed, numerous studies indicate that OCD is frequently comorbid with other disorders, including ANX, ASD, TD, major depressive disorder (MDD), and attention-deficit hyperactivity disorder (ADHD) (Gazzelone et al., 2016; Lee et al., 2019; Yang et al., 2021).

Although etiology of OC traits has been investigated, their pathophysiology remains unclear as it involves a complex interplay of neurological abnormalities, genetic predispositions, and environmental factors. Notably, OCD has been attributed to dysfunction within cortico-striato-thalamo-cortical (CSTC) circuitry (MacMaster et al., 2008).

Several neuroimaging studies over the past 30 years have implicated both structural and functional differences in CSTC circuits between OCD patients and controls in both pediatric and adult populations. Specific brain regions of the CSTC are more active when OCD subjects report more obsessions and/or compulsions (Pauls et al., 2014). Specifically, many studies show increased anterior cingulate cortex (ACC) activity with symptom provocation (Adler et al., 2000) and normalized activity after the treatment of OCD (Perani et al., 1995). Functional imaging studies have demonstrated hyperactivity in various brain regions in individuals with OCD such as in ACC, orbitofrontal cortex (OFC), and caudate (Menzies et al., 2008; Saxena &Raunch, 2000). Stein et al. (2019) reported involvement of specific CSTC circuits in the OC pathophysiology. In particular, six circuits involving prefrontal cortex, dorsal part of caudate, OFC, nucleus accumbens (NUC), amygdala, posterior putamen, and thalamus are hyperactive during the early development of OCD and are often associated with ANX and uncertainty. Additionally, prefrontal cortex, inferior parietal lobule (IPL) is also shown to be involved in interference inhibition, action restraint, and cancellation behaviors (van den Heuvel et al., 2015; van Velzen et al., 2014; Wager et al., 2005) (Table S1).

We used a set of neuroimaging phenotypes, i.e. quantitative imaging measures, including cortical thickness (CT), surface area (SA), and subcortical volume (SV) obtained from structural Magnetic Resonance Imaging (sMRI) analysis. SMRI scans were available for a subset of the participants reported in our previous genetic study (Antonyan et al., 2025). We examined the genetic basis of OCS, and the relationship between brain structural differences and childhood OCS, informed by the Research Domain Criteria (RDoC) paradigm (Cuthbert et al., 2014). Consistent with the RDoC paradigm, we used a quantitative symptom measure of OCS: the Obsessive-Compulsive Scale of the Child Behavior Checklist Scale (CBCL-OCS) (Achenbach et al., 1983; Hudziak et al., 2006), performing genetic analyses via polygenic risk scoring (PRS) (Lewis et al. 2017; Torkamani et al., 2018) of OCS structural neuroimaging endophenotypes as dependent variables. Eight cortical imaging phenotypes were evaluated as candidate endophenotypes for associations with polygenic risk. Six regions, OFC and rostral-middle frontal cortex (RMF) amongst them, showed significant but modest associations, suggesting limited but significant genetic influence on brain structure alterations.

Furthermore, we used Mendelian randomization (MR) (Burgess et al., 2015; Smith et al., 2003) to test whether there is a causal pathway linking polygenic risk scores, sMRI changes and OCS. MR was run on the same eight regions and IPL was identified as a potential causal mediator. However, IPL was not identified as a significant polygenic association. Together, this multimodal approach provides preliminary insights into structural alterations that may underlie genetic liability and clinical expression of OCS.

This unique neuroimaging genetic study of child psychiatric outpatients with a broad range of psychopathology will enable us to integrate multi-level data to better understand the etiology and pathogenesis of OCS.

To our knowledge this is a biggest study of its kind that aims to determine the relationships between genetic variation, brain structural changes and childhood OCS.

## Methods

### Participants

Subject recruitment was carried out in different sites: 1) University of Michigan, MI, and 2) Wayne State University, MI, and 3) SickKids Hospital in Toronto, ON. For genotyping, participants between the age of 8-18 provided saliva and/or blood when available (Antonyan et al., 2025). SMRI measures were obtained for 143 individuals (ages 8-30years; 87% of participants aged 9-20, median age=16years), as a follow-up assessment after genetic and clinical data collection. Subject recruitment, inclusion and exclusion criteria are explained in Table S2 and elsewhere (Antonyan et al., 2025). Our samples consist of only individuals of European ancestry.

Diagnostic specifications of OCD, ANX, ADHD, MDD, ASD, and TD that are explained in Table S3.

### Phenotypic Data

This study includes one psychiatric phenotype of OCS. OCS is measured using a subset of 8 items from the CBCL-OCS questionnaire in the range of 0-16 which is a well validated parent report standardized questionnaire on which the participant is rated on various behavioral and emotional problems that they exhibited over previous six months (Achenbach & Craig, 1983; Hudziak et al., 2006).

### Image Acquisition

All MRI acquisitions were conducted on a Siemens 3T Verio system with a 12-channel head-coil. A T1-weighted structural image was obtained utilizing a 3D Magnetization Prepared Rapid Gradient Echo (MPRAGE) sequence with the following parameters: TR: 2,150ms; TI: 1100ms; TE: 3.53ms; FOV: 256x256 mm2; matrix dimensions: 256x256; flip angle: 8; phase encoding: R/L; 160 axial slices, 1.0 mm thickness; acquisition time: 4:59. All scans were reviewed by a neuroradiologist to exclude clinically significant irregularities.

### Structural Neuroimaging Phenotypes

All 143 samples have the following sMRI measurements: CT and SA of 33 cortical regions averaged across right and left hemispheres; SV measurements of 11 limbic regions averaged across left and right hemispheres.

Overall, 77 neuroimaging phenotypes (Table S4) were collected after analyzing, editing, and obtaining the measurements from FreeSurfer software (version 7.2.0) via the “recon-all” pipeline (Dale et al., 1999; Desikan et al., 2006; Fischl and Dale, 2000). The imaging measurements were defined as mean CT (mm), mean SA (mm^2^), and mean SV (mm^3^) of left and right hemispheres in 33 ROIs for CT and SA, and 11 ROIs for SV (Table S4). Data cleaning and standardizing steps were conducted via R software packages (Anon et al., 2004) and IBM SPSS Statistics, Version 28 (IBM SPSS Statistics Version 28.0. Armonk, NY: IBM Corp.). In order to filter the phenotypes two-step prioritization was carried out: (1) discovery-based and (2) hypothesis-based.

1. *The discovery-based prioritization* was based on the strong correlation between sMRI estimates and CBCL-OCS within our sample. The strong correlations were identified via partial (Spearman) correlation with non-parametric variables controlled for biological sex and age. Additionally, false discovery rate (FDR) analysis using Benjamini-Hochberg approach was conducted to correct for multiple testing. The statistical testing was conducted using IBM SPSS Statistics, Version 28 (IBM SPSS Statistics Version 28.0. Armonk, NY: IBM Corp.).
2. *The hypothesis-based prioritization* was centered on available peer-reviewed literature and the available neuroimaging ENIGMA literature (Grasby et al., 2020; Satizabal et al., 2019). The literature search was done based on literature published on OCD and the brain regions associated with it. Only peer-reviewed articles in English were included, prioritizing publications from recent years. MEDLINE/PubMed resources were utilized for literature search purposes (Motschall & Falck-Ytter, 2005; Sood and Ghosh, 2006). Additionally, the regions with the strongest genetic disposition based on ENIGMA studies for cortical regions (Grasby et al., 2020), for subcortical regions (Satizabal et al. 2019) and for neuroimaging OCD studies (Boedhoe et al., 2018) were prioritized.

### Genotypic Data

143 samples had relevant genotyped data that was cleaned and standardized according to standard a genome-wide association study (GWAS) QC steps for further analysis. As mentioned above this is a subset of a bigger genetic study and more detailed information about genotyping methods is described elsewhere (Box S1) (Antonyan et al., 2025). Approximately 1.7 million unique single nucleotide polymorphisms (SNPs) were tested. Later imputation analysis increased this number up to 6 million SNPs. Due to low power given the small sample size, association analysis was not performed, however, to ensure the quality of the samples, standard QC steps of GWAS were carried out as explained elsewhere (Box S1) (Turner et al., 2011).

### Polygenic Risk Score Analysis

PRS analysis, which combines information from multiple SNPs (Lewis et al., 2017; Torkamani et al., 2018) was run on pre-selected neuroimaging candidate endophenotypes separately using the current samples as target data and summary statistics from ENIGMA data as discovery data.

The base data summary statistics were obtained from two ENIGMA studies (Thompson et al. 2014) and the current study is not a part of any of those studies: (1) CT and SA measurements were obtained from the study by Grasby et al. (2020) and included around 34,000 samples and around 370 genome-wide significant loci; (2) SV estimates were acquired from the study by Satizabal et al. (2019) and included around 39,000 samples and over 200 genome-wide significant loci. Both studies and our target data are based on European ancestry populations and have matching imaging phenotypes labelled by FreeSurfer (Dale et al. 1999; Fischl and Dale 2000) (Table S4).

Based on our research question a standard PRSice2 clumping+thresholding (c+t) and LDpred2 Bayesian methods were selected. Both methods have been widely used and have their challenges and limitations which we addressed carefully. More detailed methodology and limitations are explained elsewhere (Choi et al. 2019; Vilhjálmsson et al. 2015). PRSice2 was utilized as the first step as it is less computationally consuming to apply on all pre-selected phenotypes (i.e., all the imaging phenotypes that passed the two-step phenotype prioritization). Biological sex, age, age squared and the first four components from population stratification were covariates. For validation purposes permutation analysis (n=1000) was implemented (Choi et al. 2019; Choi et al., 2020). Later, LDpred2-auto was conducted as the second step on the subset of selected imaging phenotypes that had significant empirical p-value (strict P_empirical_< 0.05; loose P_empirical_ < 0.1) and significant PRS model fit (p-value < 0.05) based on PRSice2 results. The Linkage disequilibrium (LD) matrix from the HapMap3+ reference genome was used. LDpred2-auto automatically estimates sparsity P and heritability estimates (LDSC), therefore a validation set is not required. The *bigsnpr* R package (Prive et al. 2018) was used to run LDpred2 using the adapted script provided by Privé et al. (2020).

To summarize, PRS analysis allowed us to identify the combined effect of high-risk SNPs that contribute to alteration for structural changes in the pre-selected brain regions.

### Mendelian Randomization

MR analysis was carried out to determine if there is any causal interference between identified polygenic risk loci and OCS where neuroimaging measurements are the candidate endophenotypes. Instrumental variables (IV) were the SNPs with high polygenic risk for structural brain changes, structural brain changes were exposures, and OCS was the outcome. Confounders were biological sex, ethnicity (PC1-PC4), and age. More detailed explanation on MR methodology is in Fig S1. There are two widely utilized approaches of two-sample MR: inverse-variance weighted (IVW) and MR-Egger. Both methods work similarly and estimate the proportion of the variance explained by the instrumental variables (IVs), in this case genetic risk variants. We selected MR-Egger as the estimate is also sensitive to the variability between the genetic associations with the risk factor. Moreover, MR-Egger can be utilized when a stronger instrument strength independent of direct effect (InSIDE) assumption does not apply (Bowden et al. 2015; Burgess et al. 2017). InSIDE assumes that the association between genetic IVs and exposure is unrelated to the pleiotropy path from genetic IVs to the outcome that is independent exposure of interest (Burgess & Thompson, 2015; Davey Smith et al., 2003). MR-Egger tests were conducted using the *MendelianRandomization* R package (Yavorska et al., 2017). As a standard practice, the most influential observations were detected using Cook’s distance and removed via leave-one-out approach (Burgess, Bowden et al 2017; Cook, 1977). Cochran’s Q statistics was calculated to test for IVs heterogeneity bias (Burgess et al 2017). Additionally, MR-PRESSO was applied to test the validity of significant MR test and to check for horizontal pleiotropy bias (Hemani et al 2018; Verbanck et al 2018).

## Results

### Samples

143 samples were genotyped. 10 out of those individuals did not have OCS scores. Therefore, 133 samples were included for further analysis. After additional QC steps (Box S1) (Antonyan et al., 2025), 113 samples remained for PRS and MR analyses. All 113 individuals had all three levels of information: (1) neuroimaging: sMRI measurements; (2) genetic: genome-wide SNP data; (3) clinical assessment: CBCL-OCS scores.

### Phenotypic Data

77 imaging phenotypes were obtained based on FreeSurfer software labeling (Table S4) (Dale et al. 1999). The result of two-step phenotype prioritization are as follows: (1) eight regions of interest (ROI) were selected after discovery-based prioritization (Table 1) (Table S5); (2) hypothesis-based prioritization identified 11 ROIs (Table 2). We then combined the ROIs resulting in 19 pre-selected phenotypes to run the PRS analysis.

**Table 1:** Discovery-based prioritization.

|  | **sMRI** | **ROIs** | **P-value** | **FDR** |
| --- | --- | --- | --- | --- |
| 1 | SV | Amygdala | <0.001 | 0.004 |
| 2 | CT | SMG | <0.001 | 0.002 |
| 3 | CT | Cuneus | 0.002 | 0.003 |
| 4 | CT | ITG | 0.003 | 0.004 |
| 5 | CT | RMF | 0.003 | 0.004 |
| 6 | CT | PostCG | 0.003 | 0.007 |
| 7 | CT | OFC | 0.008 | 0.009 |
| 8 | CT | PreCG | 0.01 | 0.01 |

**Table 2:**
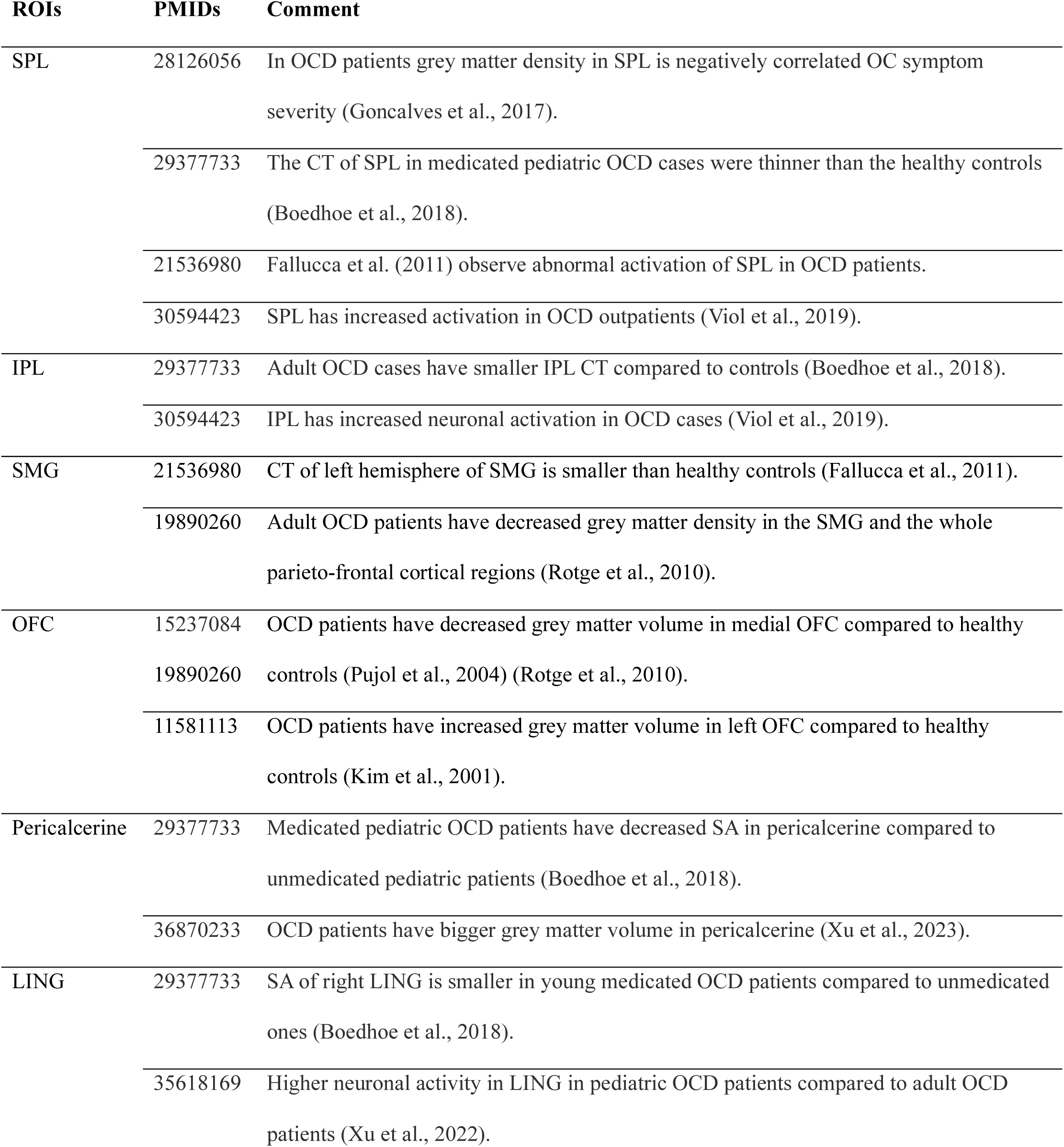
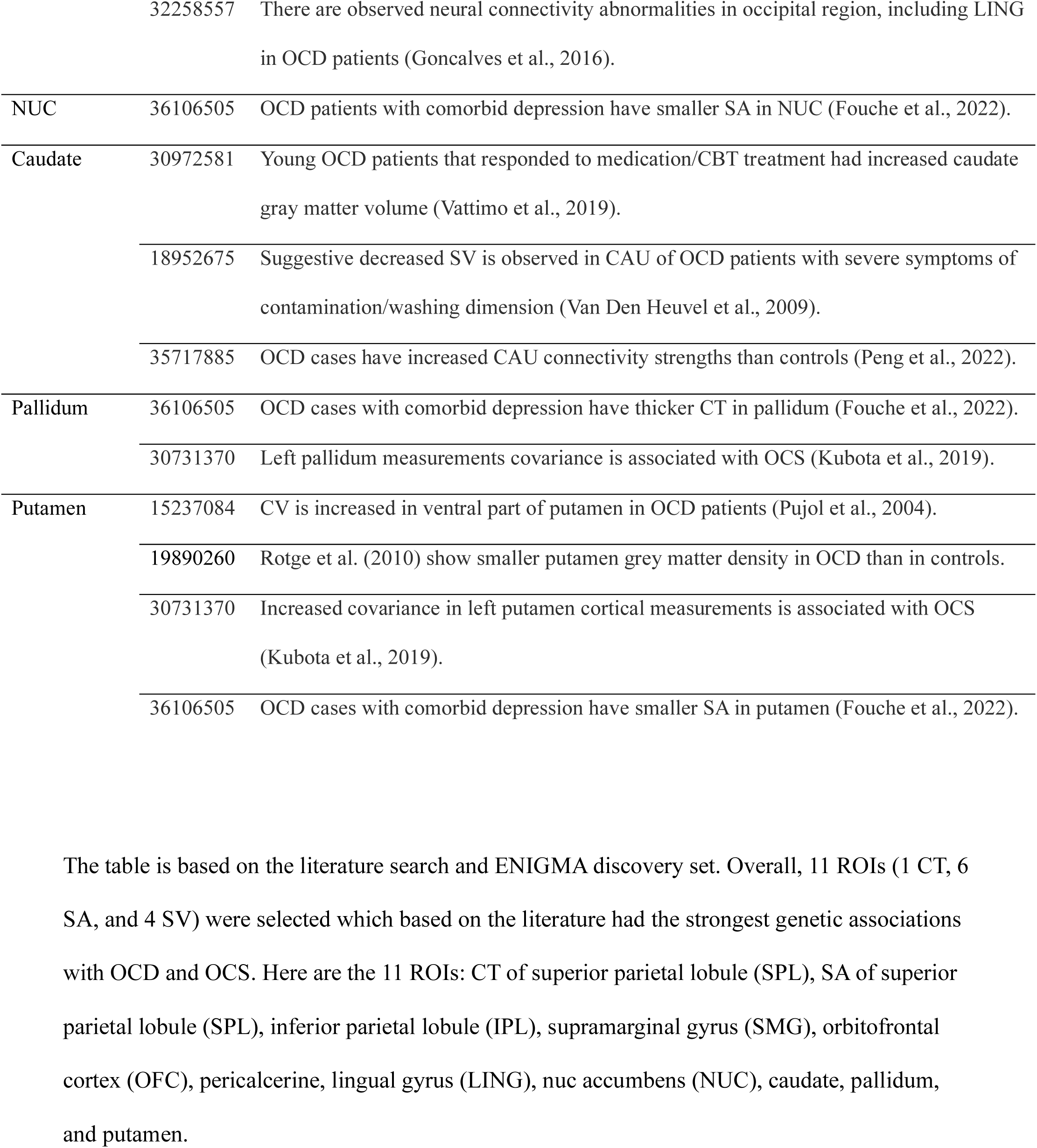
Hypothesis-based prioritization.

### Polygenic Risk Scores

PRSice2 method was carried out on all 19 ROIs and the results are reported in Table 3. LDpred2-auto was run on 8 ROIs that were selected after “hypothesis-based prioritization” and PRSice2 tests. 6 ROIs showed significant (P_Bonferroni_<0.006) but limited polygenic variance explained (Fig 1).

**Fig 1:**
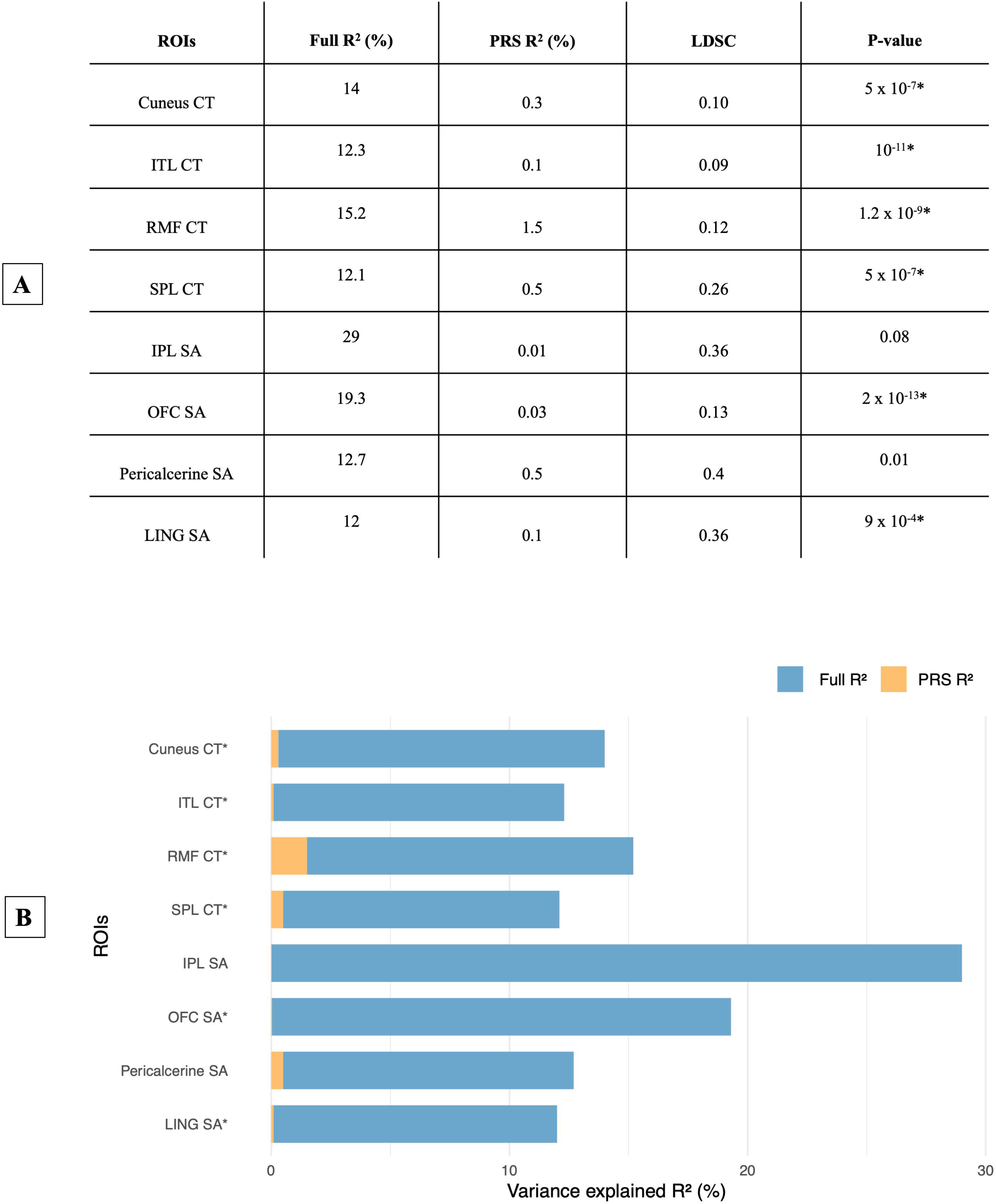
LDpred2-auto results on 8 ROIs. **A.** Full R^2^ shows the variance explained by all predictors that includes covariates and PRS; PRS R^2^ is the incremental ΔR^2^ that represents variance uniquely explained by PRS; LDSC score corresponds to heritability estimate in the analysis calculated using HapMap3+ reference data, P-value is the significance, multiple testing Bonferroni correction p-value would be 0.05/8=0.006 (8 tests). 8 tests are 5 ROIs that passed the significance of P_empirical_ after running PRSice2: CT of cuneus, RMF, and SPL, SA of IPL and LING; and 3 ROIs that had small model fit P-value (<0.05) and were also identified from *hypothesis-based prioritization*: SA of OFC, pericalcarine, CT of ITG(Table 3). Fig 1(A) shows the LDpred2-auto results of 8 ROIs. Fig 1(B) is the visual representation of the LDpred2-auto results where blue bars show the variance explained by all factors, orange bars represent the variance only explained by PRS. Abbreviations of ROIs are: ITL CT – cortical thickness of inferior temporal lobule; RMF CT – cortical thickness of rostral middle frontal; SPL CT – cortical thickness of superior parietal lobule; IPL SA-surface area of inferior parietal lobule; OFC SA – surface area of orbitofrontal cortex; LING SA – surface area of lingual gyrus.

**Table 3:**
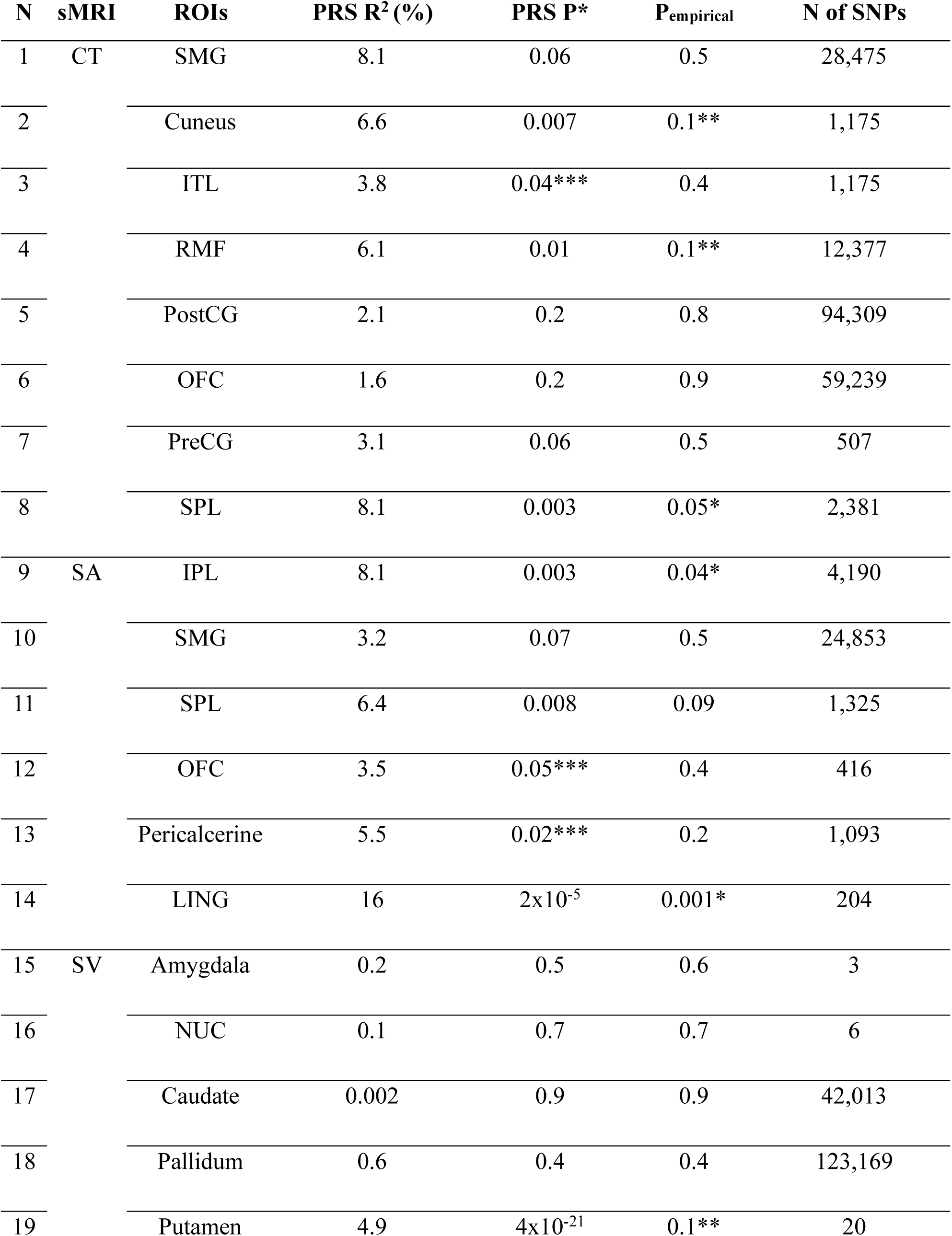
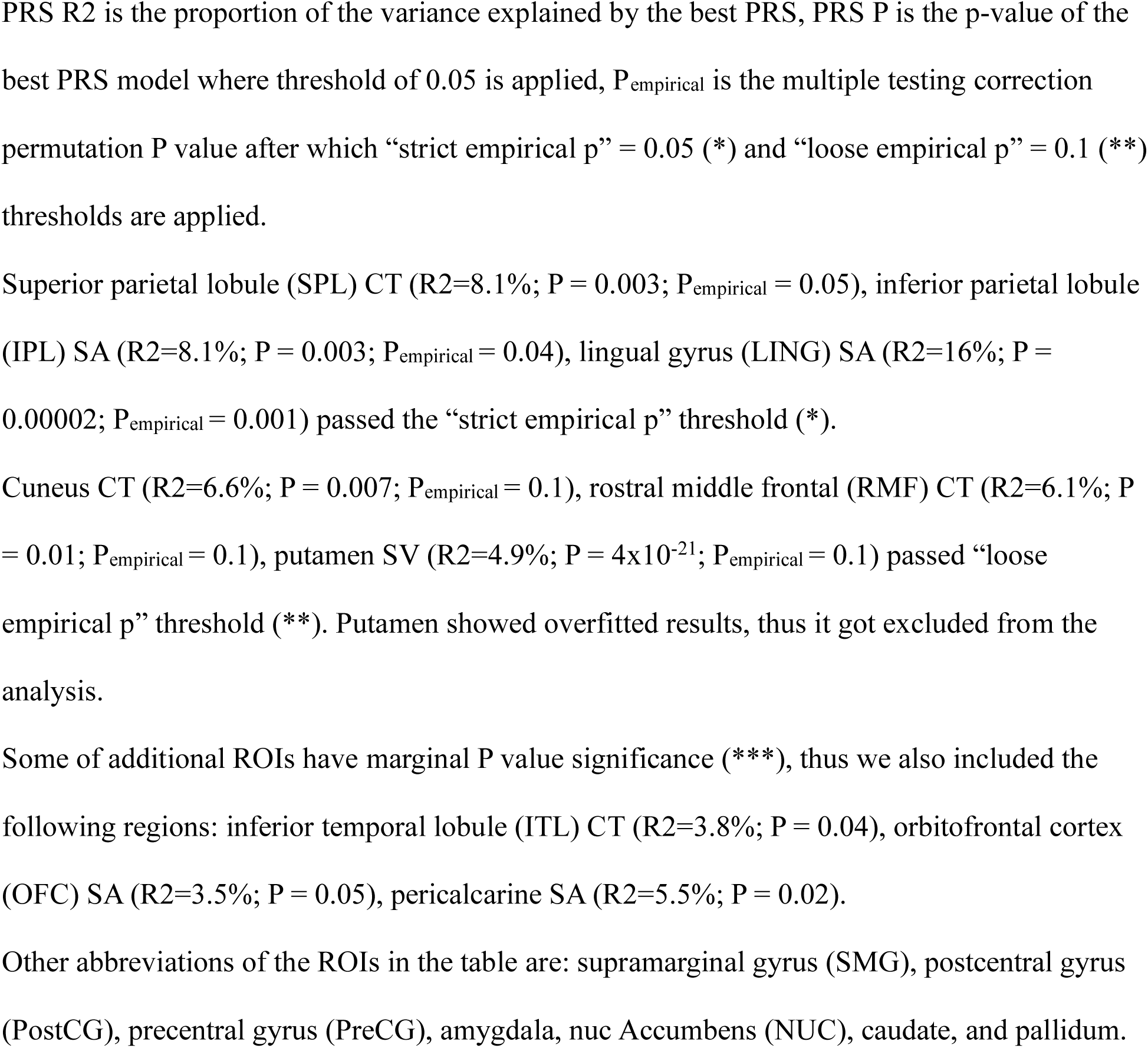
Results from PRSice2 for selected 19 phenotypes.

### Mendelian Randomization

In parallel to PRS, MR analysis via MR-Egger was performed to show if any of the correlations between genetic risk markers of the 8 ROIs show a causal interference. The SNPs that contribute to PRS were utilized as IVs in MR analysis. After Cook’s distance calculation, only IPL SA (P_Estimate_=0.03) remained significant (Table 4) (Fig S2). Moreover, no significant biases were identified for IPL SA: heterogeneity Cochran’s Q = 3104 degrees of freedom on 3671; and there was non-significant horizontal pleiotropy (p=0.8).

**Table 4:** MR-Egger Results.

| sMRI | ROIs | Estimate (SE) | P<sub>Estimate</sub> | Intercept (SE) | P<sub>Intercept</sub> | N<sub>IVs</sub> |
| --- | --- | --- | --- | --- | --- | --- |
| SA | IPL* | 0.001 (0.001) | 0.03* | -0.03 (0.01) | 0.01* | 3,673 |
|  | LING | 0.003 (0.003) | 0.3 | -0.03 (0.05) | 0.3 | 205 |
|  | Pericalcerine | 0.004 (0.003) | 0.16 | -0.04 (0.003) | 0.16 | 1,009 |
|  | OFC | 0.001 (0.000) | 0.8 | 0.001 (0.000) | 0.4 | 102,334 |
| CT | SPL | -4.9 (3.5) | 0.2 | 0.03 (0.02) | 0.09 | 2,179 |
|  | RMF | 0.97 (1.3) | 0.5 | -0.003 (0.006) | 0.7 | 12,402 |
|  | Cuneus | 0.5 (3.2) | 0.9 | 0.001 (0.02) | 0.99 | 1,134 |
|  | ITL | 0.009 (3.1) | 0.99 | 0.002 (0.2) | 0.94 | 1,179 |

Results after MR-Egger tests to show if the MR estimates are significant enough to show a causal relationship. The results are reported after running Cook’s distance. IPL (inferior parietal lobule) SA estimate was significant after Cook’s distance.

## Discussion

Although progress has been made in identifying genetic variants associated with OCS, little of the genetic variance has been explained and the neurobiological consequences of those variants are poorly understood. Here, we adopt the RDoC framework to investigate neurobiological and genetic etiology of adolescent sample. We observe structural brain changes as potential endophenotypes for OCS on 113 participants. The present analysis draws on a subset of participants from a larger cohort previously reported, where we observed the polygenic nature of OCS and their associations with six psychiatric disorders (Antonyan et al., 2025).”

The following eight imaging phenotypes, SA of IPL, lingual gyrus (LING), pericalcarine and OFC, CT of RMF, superior parietal lobule (SPL), inferior temporal lobule (ITL) and cuneus, were selected for PRS and MR analyses. SA of LING and OFC; CT of RMF, SPL, ITL, and cuneus showed modest but a significant relationship between polygenic influences on alterations in specific brain regions in OCS clinical sample. Indeed, some of the notable regions are OFC and RMF. OFC is a core region of CSTC and is shown to be of a central influence in OCS/OCD models (Menzies et al., 2008; Stein et al., 2019). RMF is not directly in CSTC circuit, but it is a part of dorsolateral prefrontal cortex which is part of dorsal cognitive CSTC and plays an crucial role in planning and emotional regulation (Stein et al., 2019).

MR analysis demonstrated a potential causal relationship involving polygenic risk variants and structural brain changes. Interestingly, we observed that only IPL SA acted as weak but significant causal exposure between PRS SNPs and OCS. IPL is in the parietal lobe and is also a part of a frontoparietal network of CSTC which is involved in maintaining cognitive and attention control, interference inhibition, action restraint and cancellation (van den Heuvel et al. 2015; van Velzen et al. 2014; Wager et al., 2005). However, in previous PRS step IPL SA did not reach significance.

The findings of this study should be considered in light of several limitations. Firstly, our sample consisted solely of participants of European ancestry, which restricts the applicability of our findings to other ethnic groups. Ongoing research efforts are aimed at increasing the diversity of samples to better represent multiple ethnic backgrounds.

A significant challenge in youth neuroimaging studies is the heterogeneity of data due to variations in the developmental stages of participants, as structural brain changes can differ based on age and development. To mitigate this issue, age and age squared (age²) were included as covariates in all statistical analyses.

A number of limitations may arise while analysing sMRI brain scans of young participants via FreeSurfer (Dale et al. 1999; Fischl and Dale 2000). It predominantly relies on adult brain templates which can lead to suboptimal segmentation when applied to developing brains. Additionally, FreeSurfer is sensitive to tissue intensity inhomogeneities in structural MRI data, which are more common at higher magnetic field strengths and can affect the accuracy of the results. All T1 images were carefully reviewed for participant motion, and any scans with motion were excluded from our analysis. FreeSurfer uses sophisticated intensity normalization and bias field correction algorithms to mitigate these and other issues such as background noise problem with 3T MRI data. however, errors can still occur, potentially impacting the reliability of surface mapping and segmentation. Thus, in some cases, manual intervention might be required.

Related to the genetic analysis, an important limitation is the inclusion of only common variants in GWAS. In future, inclusion of rare variants may enhance robustness and power of the results. Due to the small sample size, we did not perform GWAS directly on our samples. Instead, we used PRS as predictors and utilized summary statistics from large-scale ENIGMA studies to derive these scores. Additionally, MR has several integral limitations, such as the MR assumptions and selection of uncorrelated IVs (VanderWeele et al., 2014). We addressed these by checking for potential biases across all MR tests.

To confirm these findings in larger samples, further replication should focus on pediatric and adolescent individuals rather than later-onset OCD or other disorders as the early-onset OCS has higher heritability compared to those with later onset (Davis et al., 2013).

## Conclusions

To our knowledge this is the only study on adolescent samples that investigates the genetic correlation of brain changes and OCS. These findings, if they remain significant in a larger sample would provide insight to the pathophysiology of OCS, and the associated structural changes in specific brain regions that could inform the development of new prevention and treatment strategies.

## Data Availability

The T1-weighted structural image datasets generated and analyzed during the current study have been shared with the NIMH Data Archive (Collection ID: C2955).
Genomic datasets generated and analyzed during the current study are not publicly available due the fact that they consist individual-level data and are a subject of research in progress but may be available from the corresponding author on reasonable request.

## Acknowledgments

This work was part of L. Antonyan’s PhD thesis project at the University of Calgary, AB (defended December 2024). Dr. Antonyan thanks her supervisory committee members Dr. Frank MacMaster and Dr. Quan Long for their continuous support.

The authors also acknowledge the individuals involved in recruiting, screening, assessing, and scheduling research participants (Pamela F. Szura, Julia Bellamy, Usha Rajan, Shannon M. Harbin, Jenna K. Nienhuis, Caroline L. Carter, Leslie E. Kibbe, Yona E. Isaacs, Barbara S. Hanna, Haley E. Rough, and Kelsey M. Collins) for this data set. Another appreciation goes to the undergraduate students (Roy Han and Maya Antonyan) who helped with cleaning the data and required literature review.

## Author Contribution

Author contributions included conception and study design (LA and PDA), data collection, acquisition, or analysis (SS, CLB, GB, RN, FM, GLH, PE, DR and PDA), statistical analysis (LA and PE), interpretation of results (LA and PDA), drafting the manuscript work (LA) or revising it critically for important intellectual content (LA, FM, PE, and PDA) and approval of final version to be published and agreement to be accountable for the integrity and accuracy of all aspects of the work (All authors).

## Ethics approval

This study was covered by IRISS Ethics Certificate number REB16-1403_REN8. To participate in the study written informed consent and assent were obtained from parents/guardians and participants, respectively; participants over the age of 18 provided written informed consent by themselves. The Human Subjects Investigative committees at Wayne State University, the University of Michigan and the Conjoint Health Research Ethics Board at the University of Calgary approved the protocol and all methods.

## Availability of data and material

The T1-weighted structural image datasets generated and analyzed during the current study have been shared with the NIMH Data Archive (Collection ID: C2955). However, genomic datasets generated and analyzed during the current study are not publicly available due the fact that they consist individual-level data and are a subject of research in progress but may be available from the corresponding author on reasonable request.

