## Supplementary Materials for "Association of Brain Structural Measurements and Polygenic Risk Scores with Obsessive-Compulsive Symptoms in Adolescents Diagnosed with Obsessive-Compulsive Disorder, Attention-Deficit/Hyperactivity Disorder, Anxiety, Depression, Autism and Tic Disorders"

### Supplementary Material

Table S1: CSTC circuitry.

CSTC regions alterations reported for each CSTC network according to Stein et al. (2019) and van den Heuvel (2016). (1) Dorsal cognitive CSTC: pre-supplementary motor area, dorsolateral prefrontal cortex, dorsomedial prefrontal cortex, dorsal part of caudate and thalamus. (2) Ventral cognitive CSTC: inferior frontal gyrus, ventrolateral prefrontal cortex, ventral part of caudate and thalamus. (3) Ventral reward CSTC: orbitofrontal cortex (OFC), nucleus accumbens (NUC) and thalamus. (4) Frontolimbic CSTC: ventromedial prefrontal cortex, amygdala and thalamus. Additionally, dorsal cognitive CSTC and ventral cognitive CSTC with (5) sensorimotor CSTC (supplementary motor area, posterior putamen, thalamus), are active in later phases of OCD and are related to habitual behaviors (Stein et al., 2019). Alterations in (6) frontoparietal network CSTC (prefrontal cortex, inferior parietal lobule (IPL)) are involved in interference inhibition, action restraint, and cancellation behaviors (van den Heuvel et al., 2015; van Velzen et al., 2014; Wager et al., 2005). Subcortical regions such as ventral part of caudate, NUC, posterior putamen and thalamus are related to goal-directed behavior and motor control (Stein et al., 2019; van den Heuvel et al., 2015).

| **Circuit** | **Regions** | **OC related function and behavior** |
|---|---|---|
| Dorsal cognitive CSTC | pre-supplementary motor area, dorsolateral prefrontal cortex, dorsomedial prefrontal cortex, dorsal part of caudate, thalamus | planning, emotion regulation, |
| Ventral cognitive CSTC | inferior frontal gyrus, ventrolateral prefrontal cortex, ventral part of caudate, thalamus | response inhibition, goal directed behavior |
| Sensorimotor CSTC | supplementary motor area, posterior putamen, thalamus | stimulus-response based habitual behavior |
| Frontolimbic circuit | ventromedial prefrontal cortex, amygdala, thalamus | fear extinction, anxiety, uncertainty |
| frontoparietal network | Prefrontal cortex<br>Inferior parietal lobe | coordination of cognitive control, compulsions |

Table S2: The inclusion and exclusion criteria for participant recruitment.

| Inclusion Criteria | Exclusion Criteria |
|---|---|
| 1) 8-18 years of age (genotyping, clinical assessment); | 1) neurological abnormalities such as seizure disorders; |
| 2) 8- 25 (follow-up imaging collection); | 2) history of head injury with a sustained loss of consciousness; |
| 3) ambulation; | 3) chronic medical illness or known Mendelian disorder; |
| 4) good physical health; | 4) history of substance abuse or dependence; |
| 5) ability to give written informed consent (to be interviewed in English); | 5) use of psychotropic medication during the past month; |
| 6) intelligence quotient >80; | 6) recent weight loss leading to body weight less than 85% than expected for age and height; |
| 7) normal or corrected-to-normal vision. | 7) adoption; |
| | 8) use of a stimulant medication during the past 24 hours or use of a psychotropic medication other than a stimulant or selective serotonin reuptake inhibitor during the past 2 weeks. |

Table S3: Mental disorders DSM-5 and K-SADS-PL diagnostic characterizations

| | |
|---|---|
| **OCD** | Obsessive-compulsive disorder:<br>OCD cases have current Children's Yale Brown Obsessive Compulsive Scale (CY-BOCS) score ≥10 |
| **ANX** | Cases with Anxiety disorders include separation anxiety disorder, specific phobia, social anxiety disorder (social phobia), panic disorder, agoraphobia, and generalized anxiety disorder |
| **MDD** | Depressive disorders are specified as major depressive disorder and persistent depressive disorder. |
| **ADHD** | Attention-deficit/hyperactivity disorder cases included in the study who routinely discontinue a stimulant medication during school breaks. |
| **ASD** | Autism spectrum disorder cases were assessed via 2013 version of K-SADS-PL 2013 which lacks the detail of the ADI/ADOS. Instead, the SCQ was obtained on almost every case and used to identify those with scores over 14. |
| **TD** | Tic disorders are specified as Tourette's disorder, persistent motor or vocal tic disorders, and provisional tic disorder. |

Table S4: ROI labeling

79 neuroimaging phenotype measurements from the Freesurfer software via the “recon-all” pipeline (Dale et al., 1999; Fischl and Dale 2000; Desikan et al. 2006). It includes 34 ROIs for cortical thickness (CT) and surface area (SA), and 11 ROIs for subcortical volume (SV).

| **CT and SA labels** | **SV labels** |
|---|---|
| temporal sulcus | Thalamus |
| caudal anterior cingulate gyrus | Caudate |
| caudal middle frontal gyrus | Putamen |
| cuneus | Pallidum |
| entorhinal cortex | Amygdala |
| fusiform gyrus | Hippocampus |
| inferior parietal lobule | Nuc Accumbens |
| inferior temporal cortex | LateralVentricle |
| isthmus of cingulate gyrus | Ventreal DC |
| lateral occipital cortex | InfLatVent |
| lateral orbitofrontal cortex | Cerebellum |
| lingual gyrus | |
| medial orbitofrontal gyrus | |
| middle temporal gyrus | |
| parahippocampal cortex | |
| paracentral lobule | |
| pars opercularis | |
| pars orbitalis | |
| pars triangularis | |
| pericalcarine cortex | |
| postcentral gyrus | |
| posterior cingulate cortex | |
| precentral gyrus | |
| precuneus | |
| rostral anterior cingulate cortex | |
| rostral middle frontal gyrus | |
| superior frontal gyrus | |
| superior parietal lobule | |
| superior temporal gyrus | |
| supramarginal gyrus | |
| frontal pole cortex | |
| temporal pole | |
| transverse temporal gyrus | |
| insula | |

Table S5: Discovery-based prioritization

Spearman (partial) correlation using age at scanning and biological sex as covariates. 2 SV and 16 CT measurements passed the p-value<0.05 threshold (*), no SA measurement passed the threshold. Later, only 1 SV and 7 CT phenotypes passed false discovery rate (FDR) correction (**): amygdala, supramarginal gyrus, cuneus, inferior temporal gyrus, rostral middle frontal, postcentral gyrus, lateral orbitofrontal cortex and precentral gyrus.

| N | sMRI | ROI | P-value | FDR |
|---|---|---|---|---|
| 1 | SV | Lateral Ventricle | 0.6 | 0.05 |
| 2 | | Inferior Lateral Ventricle | 0.2 | 0.03 |
| 3 | | Thalamus | 0.01* | 0.008** |
| 4 | | Caudate | 0.7 | 0.05 |
| 5 | | Putamen | 0.1 | 0.03 |
| 6 | | Pallidum | 0.05 | 0.02 |
| 7 | | Hippocampus | 0.4 | 0.04 |
| 8 | | Amygdala | <0.001* | 0.004** |
| 9 | | Nuc Accumbens | 0.05 | 0.02 |
| 10 | | Ventral DC | 0.04* | 0.02 |
| 11 | | Cerebellum | 0.2 | 0.03 |
| 12 | CT | Caudal Anterior cingulate | 0.9 | 0.05 |
| 13 | | Caudal Middle Frontal | 0.04* | 0.02 |
| 14 | | Cuneus | 0.002* | 0.003** |
| 15 | | Entorhinal | 0.03* | 0.02 |
| 16 | | Fusiform | 0.08 | 0.03 |
| 17 | | Inferior Parietal | 0.03* | 0.02 |
| 18 | | Inferior Temporal | 0.003* | 0.007** |
| 19 | | Isthmus Cingulate | 0.8 | 0.05 |
| 20 | | Lateral Occipital | 0.04* | 0.02 |
| 21 | | Lateral Orbitofrontal | 0.008* | 0.008** |
| 22 | | Lingual | 0.06 | 0.03 |
| 23 | | Medial Orbitofrontal | 0.09 | 0.03 |
| 24 | | Middle Temporal | 0.1 | 0.03 |
| 25 | | Parahippocampal | 0.2 | 0.03 |
| 26 | | Paracentral | 0.03* | 0.02 |
| 27 | | Parsopercularis | 0.04* | 0.02 |
| 28 | | Parsorbitalis | 0.8 | 0.05 |
| 29 | | Parstriangularis | 0.4 | 0.02 |
| 30 | | Pericalcerine | 0.01* | 0.01** |
| 31 | | Postcentral | 0.003* | 0.006** |
| 32 | | Posterior Cingulate | 0.8 | 0.05 |
| 33 | | Precentral | 0.01* | 0.01** |
| 34 | | Precuneus | 0.02* | 0.01 |

| | | | | |
|---|---|---|---|---|
| 35 | | Rostral Anterior Cingulate | 0.5 | 0.04 |
| 36 | | Rostral Middle Frontal | 0.003* | 0.004** |
| 37 | | Superior Frontal | 0.1 | 0.03 |
| 38 | | Superior Parietal | 0.01* | 0.01 |
| 39 | | Superior Temporal | 0.2 | 0.03 |
| 40 | | Supramarginal | 0.001* | 0.001** |
| 41 | | Frontal Pole | 0.3 | 0.04 |
| 42 | | Temporal Pole | 0.3 | 0.04 |
| 43 | | Transverse Temporal | 0.2 | 0.03 |
| 44 | | Insula | 0.3 | 0.04 |
| 45 | SA | Caudal Anterior cingulate | 0.9 | 0.5 |
| 46 | | Caudal Middle Frontal | 0.06 | 0.001 |
| 47 | | Cuneus | 0.1 | 0.006 |
| 48 | | Entorhinal | 0.6 | 0.03 |
| 49 | | Fusiform | 0.9 | 0.5 |
| 50 | | Inferior Parietal | 0.2 | 0.01 |
| 51 | | Inferior Temporal | 0.2 | 0.01 |
| 52 | | Isthmus Cingulate | 0.2 | 0.02 |
| 53 | | Lateral Occipital | 0.09 | 0.005 |
| 54 | | Lateral Orbitofrontal | 0.9 | 0.05 |
| 55 | | Lingual | 0.4 | 0.03 |
| 56 | | Medial Orbitofrontal | 0.7 | 0.03 |
| 57 | | Middle Temporal | 0.4 | 0.03 |
| 58 | | Parahippocampal | 0.3 | 0.02 |
| 59 | | Paracentral | 0.7 | 0.04 |
| 60 | | Parsopercularis | 0.7 | 0.04 |
| 61 | | Parsorbitalis | 0.2 | 0.01 |
| 62 | | Parstriangularis | 0.8 | 0.04 |
| 63 | | Pericalcerine | 0.09 | 0.004 |
| 64 | | Postcentral | 0.8 | 0.04 |
| 65 | | Posterior Cingulate | 0.5 | 0.03 |
| 66 | | Precentral | 0.4 | 0.03 |
| 67 | | Precuneus | 0.3 | 0.02 |
| 68 | | Rostral Anterior Cingulate | 0.9 | 0.05 |
| 69 | | Rostral Middle Frontal | 0.3 | 0.02 |
| 70 | | Superior Frontal | 0.9 | 0.05 |
| 71 | | Superior Parietal | 0.3 | 0.02 |
| 72 | | Superior Temporal | 0.5 | 0.03 |
| 73 | | Supramarginal | 0.09 | 0.003 |
| 74 | | Frontal Pole | 0.4 | 0.03 |
| 75 | | Temporal Pole | 0.9 | 0.05 |
| 76 | | Transverse Temporal | 0.9 | 0.05 |
| 77 | | Insula | 0.1 | 0.007 |

Figure S1: Mendelian Randomization

A) A general case of MR, where it estimates whether the correlation between exposure and outcome is causal in the presence of confounders.

B) A specific case of MR adapted to our analysis, where IVs are the SNPs with higher polygenic risk for structural brain changes, structural brain changes were exposures, and OCS was the outcome. Confounders were the covariates used in the GWAS and PRS analysis, including biological sex, ethnicity (PC1-PC4), and age.

The number of assumptions for instrumental variables (IV) should be kept, such as there should be (1) an association between IV and exposure; (2) no association between IV and confounders; and 3) no association between IV and outcome. The MR estimates are considered to show pleiotropic effect under the instrument strength independent of direct effect (InSIDE) assumption explained elsewhere (Davey Smith & Ebrahim, 2003; Burgess & Thompson, 2015).

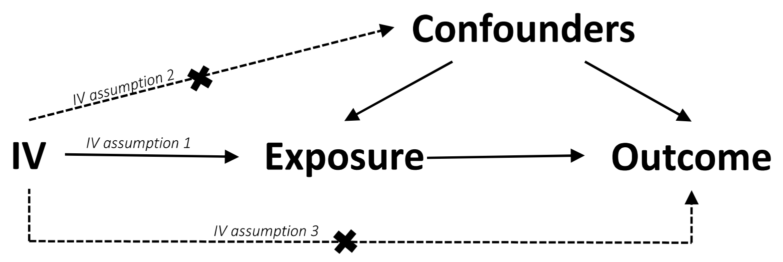


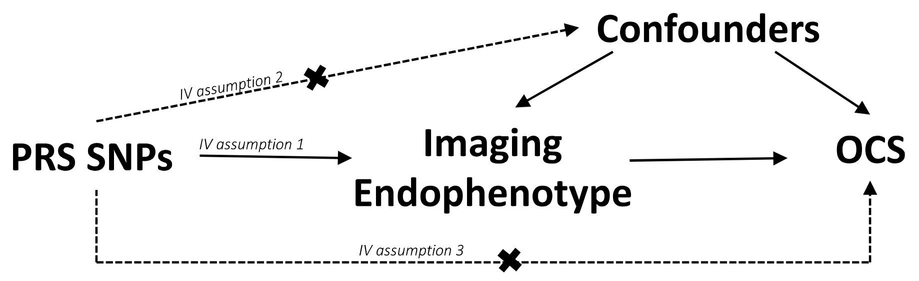

Figure S2: Inferior parietal lobule (IPL) SA estimate (0.03) was significant and has positive causal interference.

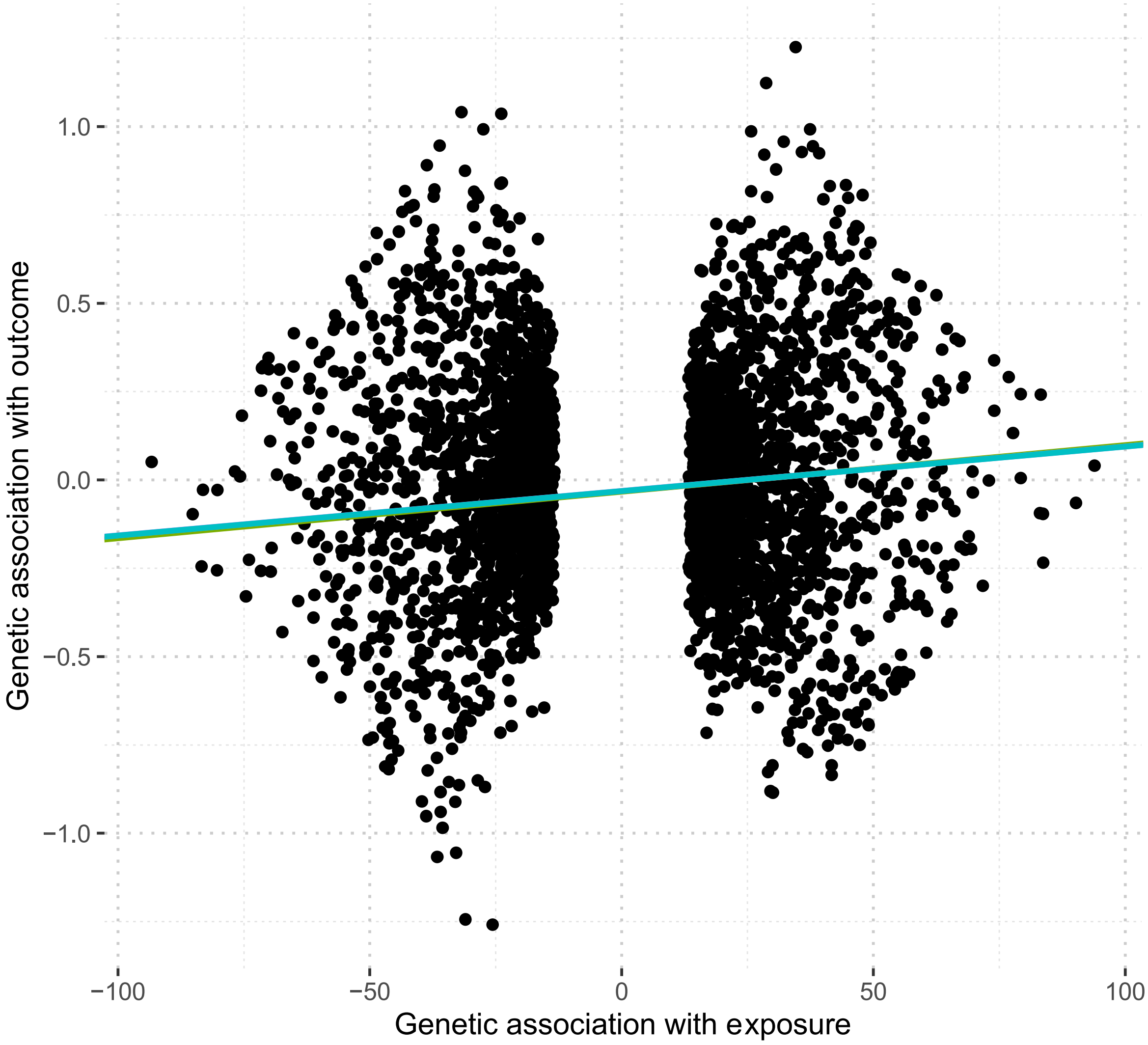
